# Digital biomarkers in early Alzheimer’s disease from wearable or portable technology: a scoping review

**DOI:** 10.1101/2025.05.02.25326845

**Authors:** Mathias Holsey Gramkow, Helena Sophia Gleerup, Anja Hviid Simonsen, Gunhild Waldemar, Kristian Steen Frederiksen

## Abstract

**Background:** The pursuit of accurate biomarkers for early detection and disease monitoring of Alzheimer’s disease (AD) has driven a growing interest in digital biomarkers. We aimed to map the research landscape of digital biomarkers in early AD obtained with wearable or portable digital health technologies (DHTs).

**Methods:** In our scoping review, we included original research on portable or wearable DHTs where digital biomarkers were measured in populations of early AD (mild cognitive impairment (MCI) or mild dementia). We searched MEDLINE, Web of Science and EMBASE with a wide search strategy with independent review and data extraction by two review team members and charted data in tabular/graphical form.

**Results:** After deduplication and screening of 8 893 records, we included 109 studies describing a wide array of wearable or portable obtained digital biomarkers. The study population consisted of 3 019 individuals with MCI due to AD (54% female, weighted mean age 73 years), and 1 942 individuals with mild AD (55% female, weighted mean age 73 years). The most studied biomarkers were rest/activity (39%), speech (17%), and gait (14%), with most studies focusing on one domain. Few studies reported outcomes associated with diagnosis (16%) and prognosis (3%).

**Conclusion:** We identified a growing evidence base investigating digital biomarkers in early AD. There is a paucity of studies examining diagnostic and prognostic properties, representing a knowledge gap. This overview may help to guide future research efforts to bridge the gap between the development and clinical implementation of digital biomarkers in early Alzheimer’s disease.

## Introduction

Alzheimer’s disease (AD) is a progressive neurodegenerative disorder characterized by cognitive decline with an insidious onset (Scheltens et al., 2016). The disease prevalence is expected to grow (Gustavsson et al., 2023), which combined with the introduction of disease-modifying treatments, will put further pressure on healthcare systems (Jönsson et al., 2023; van Dyck et al., 2023). The ongoing work for accurate and sensitive biomarkers for early detection, monitoring, and management of AD has led to a growing interest in digital biomarkers – objective, quantifiable measures obtained through various digital health technologies (DHTs) (Kourtis et al., 2019; Waldemar, 2023). These digital biomarkers could potentially be used to obtain an etiological diagnosis in areas far from specialist centres and to track the disease course and effects of pharmacological or non-pharmacological interventions (Gramkow, Waldemar, et al., 2024). While the potential of digital biomarkers is recognized (Beattie et al., 2022; Crook-Rumsey et al., 2023; Hantke et al., 2023), there is a lack of studies synthesizing knowledge on the available methods and applications.

The ability of digital biomarkers to be obtained through the use of wearable or portable technology enables use cases for remote monitoring and detection. As such, several digital health technologies, such as sensor-based technology in tablet devices and actigraphs, have been leveraged in the ongoing quest for novel biomarkers in AD (Kourtis et al., 2019). Of particular interest are the early stages of the disease, the only population currently approved for disease-modifying therapies (FDA, 2023). The early stages of AD are characterized by often subtle signs and symptoms which makes a timely diagnosis difficult (Dubois et al., 2015). Using continuous monitoring as is possible for wearable DHTs for longer periods may reveal these subtle changes. To this end, wearable technology such as actigraphy provides a means of detecting movement and circadian rhythm for an extended period in the patient’s own habitual environment (Lysen et al., 2020).

Using data collected outside the clinic setting may increase the construct validity of measurements as biomarkers are evaluated in an ecologically valid setting (Chaytor et al., 2021) and this application could be envisioned for use in early detection or treatment monitoring schemes (Hantke et al., 2023). However, knowledge of the evidence base for wearable or portably obtained digital biomarkers is scarce and this could affect research efforts, possibly limiting the harmonization of measures (Beattie et al., 2022). Only recently has the Food and Drug Administration (FDA) provided a standardized description of digital biomarkers (Vasudevan et al., 2022), which enables a further characterization of the evidence fitting this novel standard. This novel definition does not include electronic cognitive testing that only relies on computerized versions of available pen-and-paper test. The definition does however fit novel cognitive testing schemes, where sensor technology is used to measure various aspects of cognition through interaction with e.g., a tablet device (Buegler et al., 2020).

Initial results from a landscape analysis and survey of digital health technologies were recently published (Lott et al., 2024). This report focused broadly on dementia disorders, including populations with AD and it revealed a substantial catalogue of available digital health technologies that had been investigated in dementia populations. However, while technical advances are developing at a high rate, clinical implementation of digital biomarkers in memory clinics remains non-existent(Gramkow, Waldemar, et al., 2024), possibly due to barriers to implementation (van Gils et al., 2024). In this vein, there are no systematic reviews on digital biomarkers for early AD to inform clinicians of the available evidence, thus representing a knowledge gap.

This scoping review aims to comprehensively explore the landscape of research related to digital biomarkers in AD, with a specific focus on the wearable and portable digital health technologies applied to obtain them as well as their applications for clinically relevant questions such as, e.g., diagnosis, prognosis, and disease tracking. The review will shed light on the varying research applications of digital biomarkers, possibly highlighting areas lacking in research efforts from a clinical perspective. Our review will also provide data on the different digital health technologies used in early clinical AD research, whereby a possible lack of harmonization may be appreciated.

## Methods

### Study design

Systematic scoping review

### Participants, concept, and context

Studies were included if they fulfilled the following inclusion criteria: 1) explored digital biomarkers as defined by the FDA definition(Vasudevan et al., 2022), 2) applied a portable or wearable digital health technology (as evaluated by the reviewers), 3) studied the target population of adult (≥18 years old) patients with early AD (mild cognitive impairment (MCI) - mild dementia due to AD) according to the NIA-AA criteria (Albert et al., 2011; McKhann et al., 2011), 4) original research article (observational or interventional studies) written in the English language.

Studies were excluded if they fulfilled one or more of the following exclusion criteria: 1) only measures and reports on cognitive domains (e.g., tablet-administered cognitive testing for memory assessment), 2) reports solely on patients with moderate-severe dementia due to AD, 3) reports solely on patients with individuals at high risk of developing AD (i.e., amyloid-positive populations without cognitive complaints) 4) reports solely on AD in Down’s syndrome 5) qualitative studies, reviews, and conference abstracts, 5) reports solely on patient-safety, communication enhancing, and general health-related measures (i.e., non-clinical aspects), 6) reports on digital biomarkers values in ≤ 10 participants with early AD, 7) only reports on populations with mixed etiology (aggregation on dementia syndrome or mixed pathologies, such as AD with cerebrovascular co-pathology).

### Information sources and search strategy

The databases MEDLINE, Web of Science, and EMBASE were searched from inception till the 15^th^ of January 2025. Search strings including key intervention (digital biomarker/digital health technologies) search terms targeting portable/wearable DHTs were used. See Supplementary Material.

### Study records and selection process

The online software Covidence® was used for reference handling, deduplication, and screening. Titles and abstracts were screened independently by two reviewers (MHG and HSG). If disagreement occurred, a third reviewer (KSF) was consulted. Full-text articles were screened for inclusion following a similar procedure.

### Data items and charting of data

The following items were extracted from records using a customized data extraction sheet (Microsoft® Excel): 1) type of study (observational, RCT, prospective cohort study, etc.), 2) year of study, 3) number of participants with AD, 4) demographics of AD patients (age and sex), 5) disease stage (MCI, mild dementia or both), 6) setting (home, out-patient, in-patient, etc.), 7) digital health technology applied (e.g. actigraph, tablet, pupillometer, etc.) and software algorithm (if applicable), 8) digital biomarker(s) measured (e.g., rest/activity, gait kinematics, sleep, speech, pupil size, etc.), 9) outcome(s) assessed in the study (e.g., diagnosis, prognosis, disease tracking, etc.). We charted data with results presented in tabular/graphical form and performed a narrative synthesis of results. No critical appraisal of evidence was carried out.

A protocol for the review was uploaded to *figshare* (DOI: https://doi.org/10.6084/m9.figshare.24541906.v1) prior to the initiation of the study. R (ver. 4.4.2)(R Core Team, 2021) with the integrated development environment RStudio (ver. 2024.12.1) was used to create line- and bar plots using the *ggplot2* package with *ggthemr* and to calculate weighted means for the age of the diagnostic groups. The results are reported according to the Preferred Reporting Items for Systematic reviews and Meta-Analyses extension for Scoping Reviews (PRISMA-SCr) guidelines (Tricco et al., 2018) (see Supplementary Material for checklist).

## Results

### Study characteristics

After de-duplication, 8 893 references were screened for titles and abstracts and 433 records underwent full-text screening, resulting in 109 studies that were included in the review (for PRISMA flow chart see **Figure 1**). The study population totalled 3 019 individuals with MCI due to AD (55% female, weighted mean age 73 years), and 1 942 individuals with mild AD (54% female, mean age 73 years) (**Supplementary Table 1**). The studies described a wide variety of wearable or portable obtained digital biomarkers, such as rest/activity, sleep, speech and pupil size. The vast majority of studies were published within the last 10 years (85%) (**Figure 2**). The references for the included studies can be found in the **Supplementary Material**.

**Figure 1.**
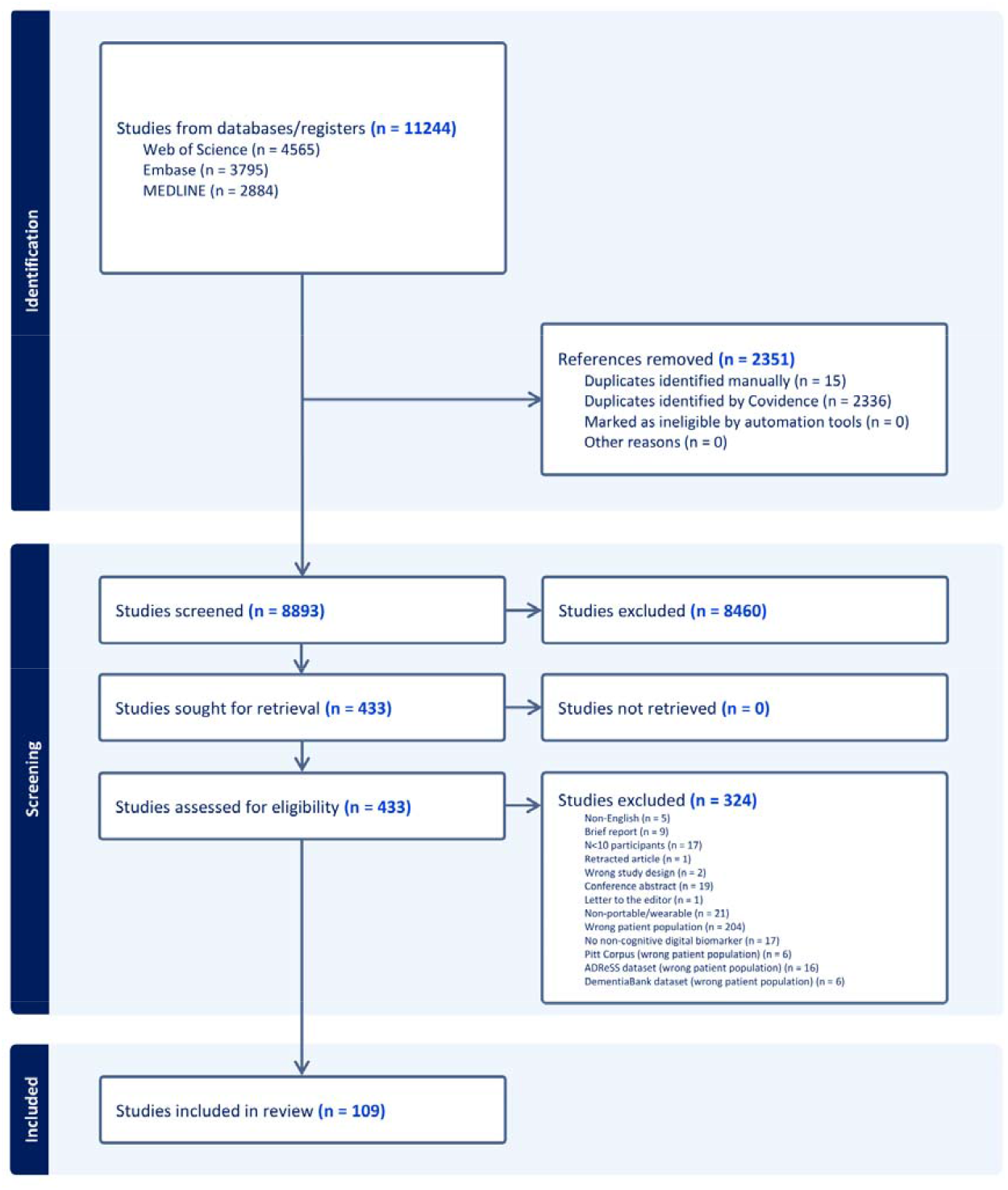
PRISMA flow chart of included studies

**Figure 2.**
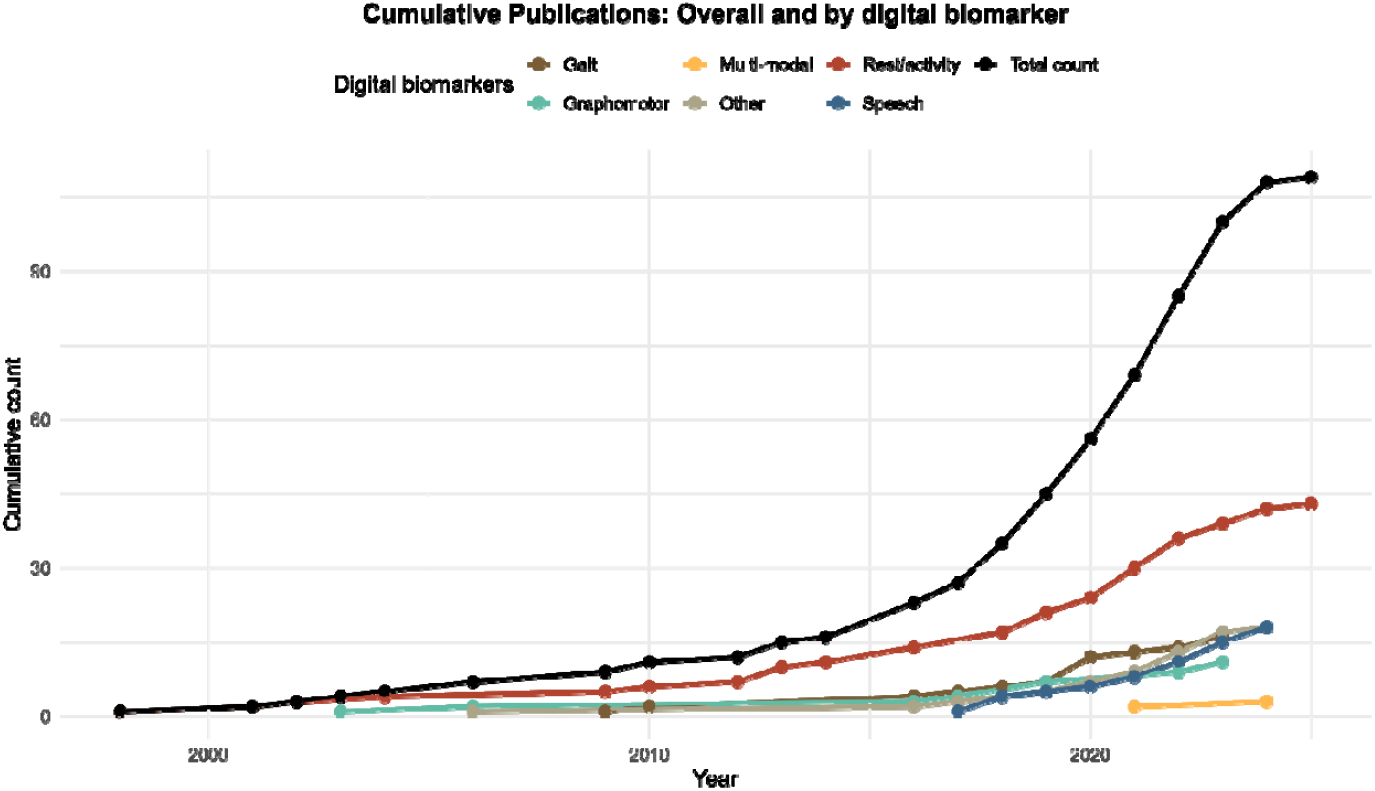
Cumulative counts of published studies on digital biomarkers from wearable or portable digital health technology in early AD (total count of publications shown for all publications in black).

### Digital biomarkers

Most studies described results on rest/activity (39%), speech (17%), gait (14%) and graphomotor function, e.g., assessing fine motor skills on a digitizing tablet device (12 %), with most studies focusing on one biomarker domain (**Figure 2**). Multi-modal evaluation of digital biomarkers was performed in three studies: Meier et al. used the Altoida application, which utilizes the internal accelerometer and touch display of a tablet device to gather several parameters during a hide-and-seek task (Meier et al., 2021); Muurling et al. explored the concurrent evaluation of sleep from both actigraphs and an EEG headband as well as continuous heart rate from the FitBit, reporting feasibility and usability aspects of this combination of markers (Muurling et al., 2024); in the study by Yamada et al., data on gait, speech and graphomotor function were collected from a motion camera and an iPad tablet to inform on classification of AD and MCI (Yamada et al., 2021).

### Digital health technologies

The most studied digital health technology was actigraphs representing 45% of studies (see **Figure 3** and **Supplementary Table 2**). Studies investigating sleep, which most often relied on actigraphic measures, included additional portable biomarkers such as wearable EEG in only one of these studies (Muurling et al., 2024). Most studies (94%) applying actigraphy used research-grade actigraphs, although manufacturers differed between studies (Supplementary Table 2). Some portable digital health technologies that were used in several studies were hand-held pupillometers (El Haj et al., 2022; Gramkow, Clemmensen, et al., 2024; Granholm et al., 2017; Kawasaki et al., 2020) to investigate pupillary responses and digitizing tablets were often used for studying graphomotor function. Studies that focused on speech biomarkers relied on both high-quality external recorders (N = 4, 24%) and built-in microphones in consumer devices such as tablets and smartphones (N = 10, 59%), where stated.

**Figure 3.**
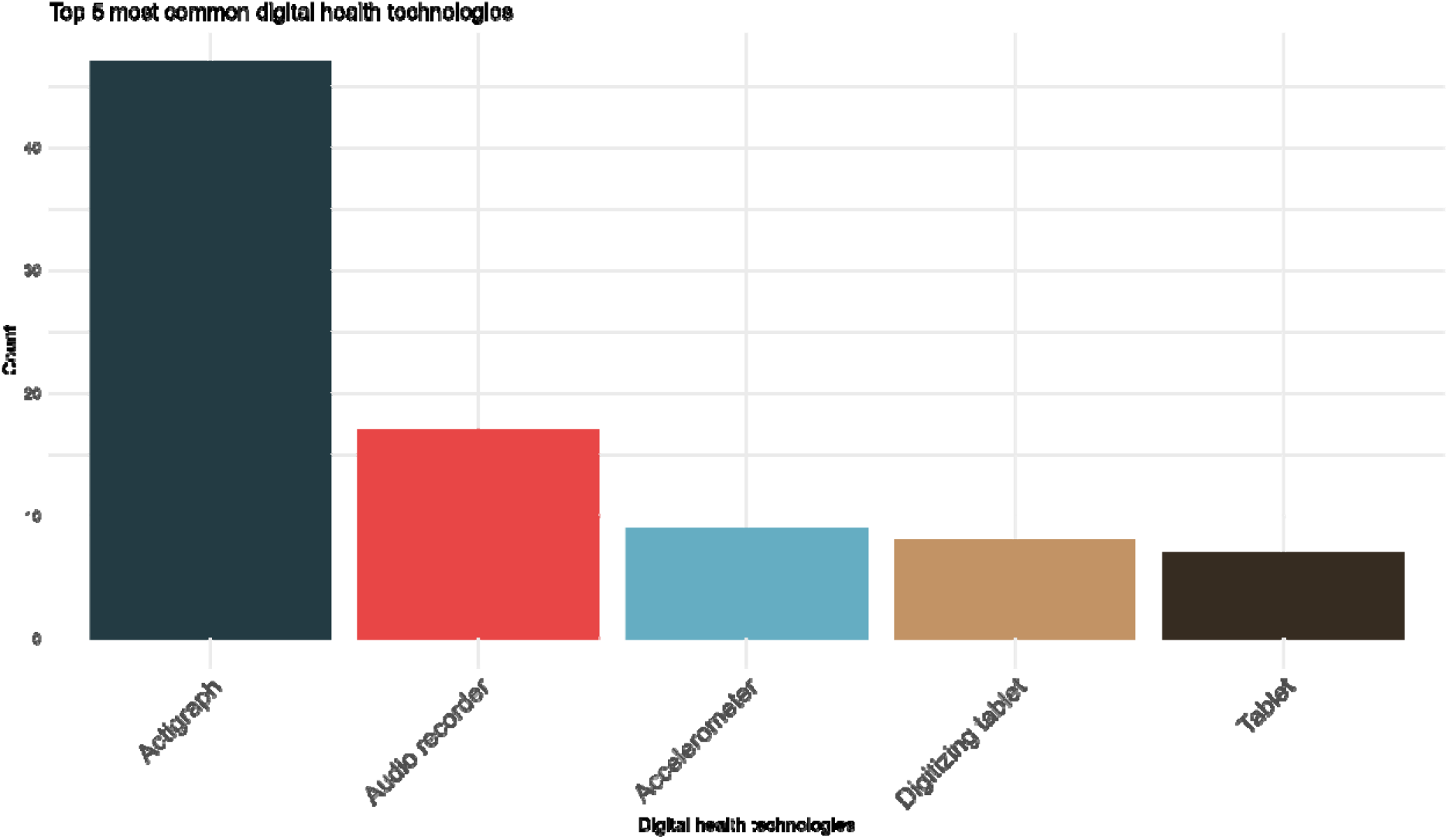
A barplot showing the number of studies applying the annotated portable or wearable digital health technologies.

### Study outcomes and setting

Most studies (72%) were descriptive in nature, comparing one or more digital biomarker measures between groups of mild AD/MCI and often a control group (**Figure 4**). Some studies reported outcomes associated with diagnosis (16%), with the majority of these studies concerning speech biomarkers (N=9, 53%). Few studies reported on a prognostic outcome (N=3, 3%), and only one study reported on a digital biomarker, namely speech, for the assessment of treatment monitoring (Hegedűs et al., 2024). There was only one study focused on feasibility aspects, in particular, that of multi-modal digital biomarker data collection (Muurling et al., 2024). A total of 70% of studies were performed in an outpatient setting, while the second most commonly included setting was community (21%), whereas 12% of studies included a home setting.

**Figure 4.**
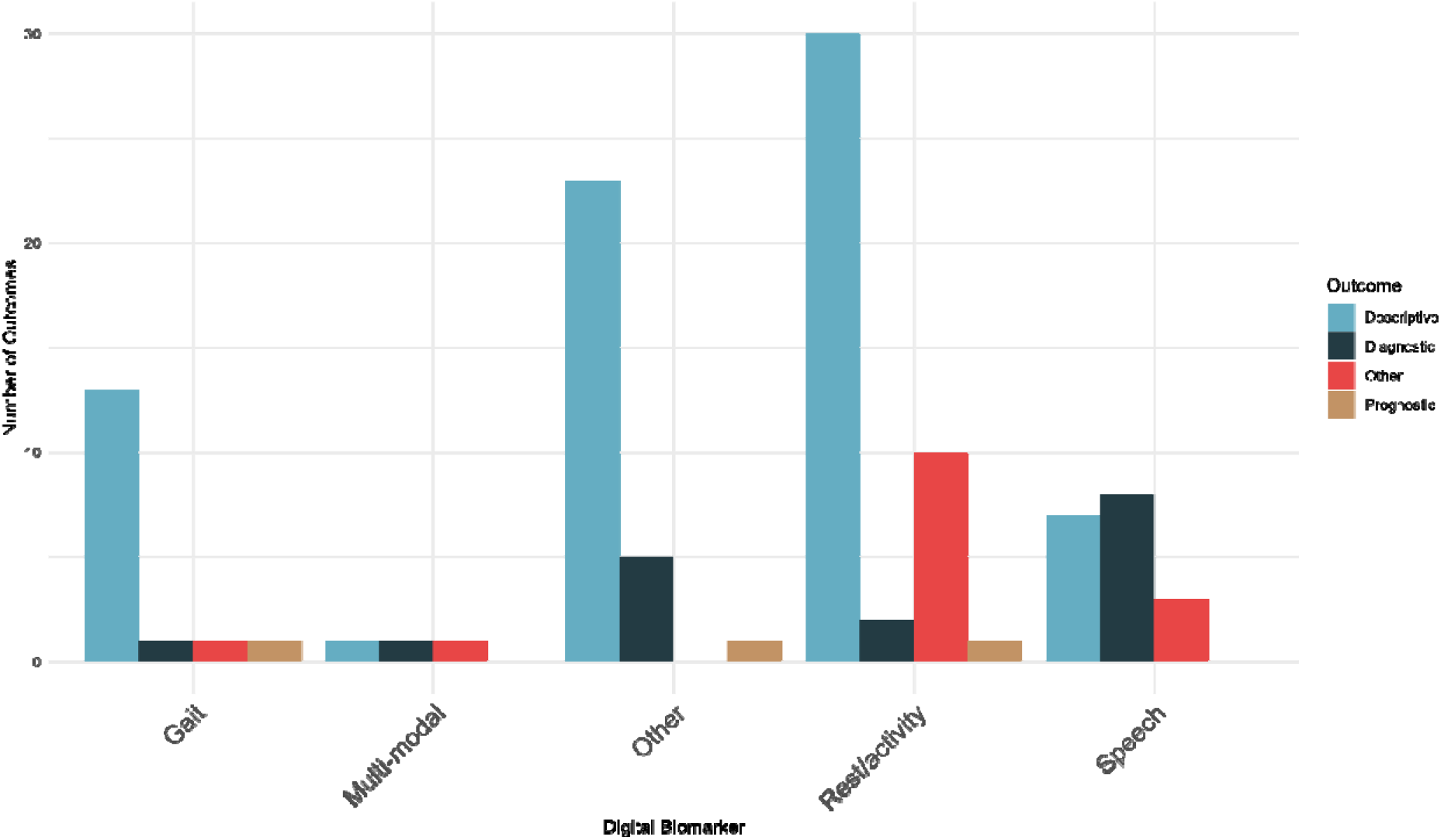
Outcomes assessed in the individual studies stratified by the digital biomarker evaluated.

## Discussion

In this scoping review, we collated available evidence on wearable and portable digital biomarkers in the early AD population using a systematic approach. We found a sizeable and growing collection of evidence, investigating digital biomarkers of which a considerable proportion of studies were focused on rest/activity. The digital health technology used to obtain evidence varied between studies, for example regarding manufacturers. Most studies investigating rest/activity, which was the most studied digital biomarker, used research-grade actigraphs. We mainly identified descriptive studies performed in an outpatient setting, while a lack of studies reporting on diagnostic, prognostic, or treatment-related measures was identified. These findings suggest a knowledge gap regarding the clinical usefulness of digital biomarkers in the early AD population, which should be the focus of future research in this area.

The initial results of a recently published landscape analysis (Lott et al., 2024) revealed a vast collection of studies that apply digital health technologies in AD and related disorders. They used the V3 framework (Goldsack et al., 2020). A framework developed by the Digital Medicine Society to promote and facilitate validation of digital health technology - to describe the validation stages of the included studies, which mention three categories of validation: verification, technical/analytical validity, and finally clinical validation. Since our review focused on studies within the clinical realm, all included studies were classified as clinical validation. This suggests that a broad range of technologies and digital biomarkers have progressed into the clinical phase of validation in early AD. In the recently updated V3+ framework (DiMe, 2025), usability aspects are now included. Usability studies are essential to guide clinicians in adopting technologies in their practice, and are equally important as clear results relating to e.g., diagnostic properties given by, e.g., sensitivity/specificity. We identified only one study by van Gils et al. (Muurling et al., 2024) focusing on the usability and feasibility of a multi-modal digital biomarker application. Usability aspects should therefore be included in future studies on digital health technology in AD to further the adoption of DHTs.

We found that diagnostic measures were most often investigated for speech, which represents a promising digital biomarker given the early affection of language in AD (Eyigoz et al., 2020). In addition, speech lends itself to easy obtainment from consumer devices, which were favorably represented amongst the DHTs used to assess speech. However, a main challenge for the field is to define minimum technical requirements for a particular mobile or tablet application for a marker such as speech to be recorded from a consumer device. An International Organization for Standardization/Technical Specifications (ISO/TS) (file number 82304-2:2021) paper describing the technical specifications to be met for health and wellness applications was published in 2021. As clinicians, selecting manufacturers that comply with this standard would increase the technical quality of the digital biomarkers that are being assessed, which will likely have an impact on relevant measures such as diagnostic performance. For speech biomarkers to move into clinical settings, the focus should be on designing and conducting studies where speech biomarkers are incorporated into the diagnostic pathways and the add-on value is investigated alongside traditional clinical scales (cognitive tests and neuropsychological evaluation) and biomarkers (neuroimaging and fluid biomarkers). A recently initiated study (Konig et al., 2023) investigating speech analysis as a screening method for inclusion in trials supports this notion.

We found a substantial number of studies investigating actigraphy as a means of obtaining data on rest, activity and sleep. Actigraphy offers a non-invasive and low-cost method for gauging activity levels and ecological behavior (Mc Carthy et al., 2016). Relatively few studies applying actigraphy reported on diagnostic properties in early AD. To translate to clinical implementation, clinicians would need to know the diagnostic value of this promising biomarker. Results from our own group show that there is diagnostic value to be gathered from actigraphy recordings in patients with early AD (unpublished data) and these results will need to be replicated in other cohorts.

Our study has several strengths. We applied a broad systematic and reproducible search strategy to capture a wide range of digital health technologies and we used an independent review process to reduce bias in the selection of studies. We also acknowledge some limitations. First, we only included studies reporting in the English language, which limited our scope. Second, we only focused on full-text published reports, which may have hindered the detection of some more recently developed DHTs. However, including only peer-reviewed reports ensures that the evidence base is validated in the field.

In conclusion, we comprehensively surveyed the available literature on applications of wearable and portable digital health technologies and digital biomarkers obtained in a population of early AD, representing a clinically relevant study cohort given the recent approval of novel disease-targeted therapies. We found a substantial amount of evidence, mainly of a descriptive nature, whereas studies investigating the diagnostic and especially the prognostic properties of digital biomarkers were uncommon. We thus identified a knowledge gap that should be bridged to bring digital biomarkers into the AD clinical realm. Given the accruing maturity of the digital field, efforts should now be focused on answering clinically relevant research questions regarding the clinical value and usability of digital biomarkers to further their adoption into practice.

## Supporting information

PRISMA-SCr checklist

## Data Availability

All data produced are available online at https://osf.io/dt7y2/

## Acknowledgments

No specific funding was obtained for this study.

## Data availability statement

The full dataset is available in the Supplementary Material.

## Code availability statement

Only elementary coding was used to produce the plots in this paper.

