## Supplementary Material for "Digital biomarkers in early Alzheimer’s disease from wearable or portable technology: a scoping review"

**Supplementary Table 1.** Descriptives of the included studies. Superscript letters identify identical cohorts.

| **First author, year** | **Study design** | **Setting** | **Outcome** | **Digital**  **biomarker(s)** | **MCI,**  **N (% female)**  **mean age** | **Mild AD,**  **N (% female)**  **mean age** | **MCI/mild AD**  **N (% female)**  **mean age** |
| --- | --- | --- | --- | --- | --- | --- | --- |
| (André et al., 2019) | Observational, cross-sectional | Home | Descriptive | Sleep | 22  - | - | - |
| (Balouch et al., 2022) | Observational, cross-sectional | Out-patient | Descriptive | Sleep | 8 (62.5%)  73.75 | 15 (33 %)  75.87 | - |
| (Basta et al., 2019) | Observational, cross-sectional | Home | Descriptive | Sleep | 73  - | 42  - | - |
| (Beltrami et al., 2018) | Observational, cross-sectional | Out-patient | Descriptive | Speech | 16  64.19 | 8  - | - |
| (Buchman et al., 2012)^a^ | Observational, longitudinal cohort study | Home | Incidence | Physical activity | - | - | - |
| (Buchman et al., 2020)^a^ | Observational, longitudinal cohort study | Home | Incidence | Gait | - | - | - |
| (Buegler et al., 2020)^b^ | Observational, longitudinal cohort study | Out-patient and home | Prognostic | Motor behavior | - | 109  - | - |
| (Callow et al., 2024)^c^ | Observational, cross-sectional | Community | Descriptive | Sleep and activity | 19 (74 %)  72.80 | - | - |
| (Callow et al., 2025)^c^ | Observational, cross-sectional | Community | Descriptive | Sleep and activity | -  74.20 | - | - |
| (Camargos et al., 2014) | Randomized controlled trial | Out-patient | Treatment efficacy | Sleep | - | 12  - | - |
| (Cavuoto et al., 2019) | Observational,  cross-sectional | Out-patient | Descriptive | Sleep | 12 (58%)  77.17 | - | - |
| (Chen et al., 2020) | Observational, longitudinal cohort study | Out-patient | Descriptive | Gait | 39 (41 %)  - | - | - |
| (Corbi & Burgos, 2022) | Observational, cross-sectional | Home | Descriptive | Sleep | - | 15 (100 %)  - | - |
| (Costa et al., 2016) | Observational, cross-sectional | Out-patient | Diagnostic | Kinematics | - | 36 (67 %)  76.00 | - |
| (Egas-López et al., 2022) | Observational, cross-sectional | Out-patient | Diagnostic | Speech | - | 25 (52 %)  72.56 | - |
| (El Haj et al., 2022) | Observational, cross-sectional | Out-patient | Descriptive | Pupil size | - | 24 (58 %)  72.33 | - |
| (García-Gutiérrez et al., 2023) | Observational, cross-sectional | Out-patient | Amyloid status | Speech | 52 (65 %)  73.80 | - | - |
| (García-Gutiérrez et al., 2024) | Observational, cross-sectional | Out-patient | Diagnostic | Speech | 826 (62 %)  74.40 | - | - |
| (Ghosal et al., 2022)^d^ | Observational, cross-sectional | Home | Descriptive | Physical activity | - | - | - |
| (Gillain et al., 2016) | Observational, longitudinal cohort | Out-patient | Descriptive, prognostic | Gait kinematics | 13 (46 %)  73.07 | - | - |
| (Gillain et al., 2009) | Observational, cross-sectional | Out-patient | Descriptive | Gait kinematics | 14 (21 %)  72.85 | - | - |
| (Gosztolya et al., 2019) | Observational, cross-sectional | Out-patient | Diagnostic | Speech | - | 25  73.96 | - |
| (Gramkow et al., 2024) | Observational, cross-sectional | Out-patient | Diagnostic | Pupil size | 28  - | 75  - | - |
| (Granholm et al., 2017) | Observational, cross-sectional cohort study | Community | Descriptive | Pupil size | 53 (0 %)  61.76 | - | - |
| (Guarnieri et al., 2020) | Observational, cross-sectional | Out-patient | Diagnostic | Rest-activity and sleep | 28 (43 %)  80.57 | 42 (62 %)  75.55 | - |
| (Hajjar et al., 2023) | Observational, cross-sectional | Out-patient | Diagnostic | Speech | 114 (57 %)  64.90 | - | - |
| (Harms et al., 2022)^b^ | Observational, cross-sectional | Out-patient | Descriptive and diagnostic | Motor behavior | - | - | - |
| (Hatfield et al., 2004) | Observational, cross-sectional, and longitudinal cohort study | Out-patient | Descriptive | Rest/activity | - | 13  67.30 | - |
| (Hegedűs et al., 2024) | Intervention, open-labeled, prospective cohort study | Out-patient | Treatment monitoring | Speech | - | 20 (70 %)  73.75 | - |
| (Huang et al., 2022) | Observational, cross-sectional | Out-patient | Descriptive | Gait kinematics | 37  - | 11  - | 48 (56 %)  65.70 |
| (Hu et al., 2009) | Observational, cross-sectional | Home | Descriptive | Rest-activity | - | 13 (31 %)  68.50 | - |
| (Ishikawa et al., 2019) | Observational, cross-sectional | Out-patient | Descriptive | Graphomotor | 25 (60 %)  75.90 | - | - |
| (Iwata et al., 2013) | Observational, cross-sectional | Home | Descriptive | Physical activity | 10 (40 %)  73.29 | - | - |
| (Jeon et al., 2023) | Observational, cross-sectional | Out-patient | Descriptive | Gait kinematics | 68 (32 %)  76.10 | - | - |
| (Kaneshwaran et al., 2019)^a^ | Observational, cross-sectional | Home | Descriptive | Sleep | - | - | - |
| (Kawasaki et al., 2020) | Observational, cross-sectional | Out-patient | Descriptive | Pupil size | - | - | 16 (63 %)  77.20 |
| (Khosroazad et al., 2023) | Observational, cross-sectional | Out-patient | Descriptive | Sleep | 50 (64 %)  74.28 | - | - |
| (Khou et al., 2018) | Observational, cross-sectional | Out-patient | Descriptive | Sleep | - | 35 (31 %)  73.66 | - |
| (Kim et al., 2021) | Randomized controlled trial | Out-patient | Treatment efficacy | Sleep | - | 22  - | - |
| (Kimura et al., 2023) | Observational, longitudinal cohort study | Out-patient | Descriptive | Sleep and activity | 118 (56 %)  75.70 | - | - |
| (Kobayashi et al., 2022) | Observational, cross-sectional | Out-patient | Descriptive | Graphomotor | 65 (42 %)  74.50 | - | - |
| (König et al., 2017) | Observational, cross-sectional | Out-patient | Descriptive | Gait | 24 (67 %)  75.00 | - | - |
| (Konig et al., 2017) | Observational, cross-sectional | Out-patient | Diagnostic | Speech | 44 (57 %)  76.30 | - | - |
| (Kuhlmei et al., 2013) | Observational, cross-sectional | Institution and home | Descriptive | Activity | 21 (95 %)  82.70 | - | - |
| (Lassila et al., 2018) | Observational, cross-sectional | Out-patient | Descriptive | Blood pressure and heart rate | 53 (68 %)  75.10 | - | - |
| (Lazarou et al., 2023) | Intervention study | Home | Descriptive | EEG frequency analysis | 13 (69 %)  72.08 | - | - |
| (Leger et al., 2016) | Randomized controlled trial | Out-patient | Descriptive | Sleep | - | 144  - | - |
| (K. Li et al., 2023) | Observational, cross-sectional | Out-patient | Descriptive | Graphomotor function | 30 (43 %)  69.23 | - | - |
| (Lim et al., 2013)^a^ | Observational, longitudinal cohort study | Community | Diagnostic | Sleep | - | - | - |
| (P. Li et al., 2020)^a^ | Observational, longitudinal cohort study | Community | Prognostic | Rest/activity | - | - | - |
| (P. Li et al., 2023)^a^ | Observational, longitudinal cohort study | Community | Descriptive and incidence | Sleep | - | - | - |
| (P. Li et al., 2018)^a^ | Observational, longitudinal cohort study | Community | Incidence | Rest/activity | - | 564  - | - |
| (Louzada et al., 2022) | Randomized controlled trial | Out-patient | Symptom monitoring | Sleep | - | 10  - | - |
| (Luboshitzky et al., 2001) | Observational, cross-sectional study | Out-patient | Descriptive | Sleep | - | 12  - | - |
| (Lu et al., 2018)^e^ | Observational, cross-sectional cohort study | Community and out-patient | Descriptive | Activity | - | - | - |
| (Lu et al., 2024)^e^ | Observational, longitudinal cohort study | Community | Descriptive and incidence | Activity | 130 (51 %)  81.50 | - | - |
| (Lysen et al., 2020) | Observational, longitudinal cohort study | Community | Incidence | Activity | - | 47  - | - |
| (Maquet et al., 2010) | Observational, cross-sectional study | Out-patient | Descriptive | Gait | 14 (50 %)  73.00 | 6 (50 %)  74.00 | - |
| (Mc Ardle et al., 2023)^f^ | Observational, cross-sectional | Community | Descriptive | Gait | 18  - | - | - |
| (Mc Ardle, Del Din, Donaghy, et al., 2020)^f^ | Observational, cross-sectional | Community | Descriptive | Walking activity | - | - | - |
| (Mc Ardle, Del Din, Galna, et al., 2020)^f^ | Observational, cross-sectional study | Community | Diagnostic | Gait | - | - | - |
| (Ardle et al., 2018)^f^ | Observational, cross-sectional | Out-patient | Descriptive | Gait | - | 17 (47 %)  67.41 | - |
| (Meier et al., 2021)^b^ | Observational, longitudinal cohort study | Out-patient | Descriptive | Multi-modal | 213  - | - | - |
| (Meilan et al., 2017) | Observational, cross-sectional | Out-patient | Descriptive | Speech | 38 (74 %)  75.61 | - | - |
| (Moghadami et al., 2021) | Observational, cross-sectional | Out-patient | Descriptive | Eye-tracking (fixation) | - | 19 (58 %)  59.00 | - |
| (Müller et al., 2017) | Observational, cross-sectional | Out-patient | Descriptive | Graphomotor | 30 (50 %)  65.30 | 20 (55 %)  69.60 | - |
| (Muurling et al., 2020) | Observational, cross-sectional | Out-patient | Descriptive | Gait | 58 (38 %)  71.00 | - | - |
| (Muurling et al., 2024) | Observational, longitudinal cohort study | Out-patient | Feasibility | Activity, sleep, heart rate | 65 (42 %)  70.00 | - | - |
| (Nasrolahzadeh et al., 2018) | Observational, cross-sectional | Community | Descriptive | Speech | - | 30 (47 %)  73.18 | - |
| (Park et al., 2021) | Observational, cross-sectional study | Community | Descriptive | Activity | 25 (52 %)  75.00 | - | - |
| (Perez-Valero et al., 2022) | Observational, cross-sectional | Out-patient | Descriptive | Electrocortical activity | - | 11 (64 %)  68.30 | - |
| (Petrillo et al., 2023)^g^ | Observational, cross-sectional | Out-patient | Descriptive | Kinematics | - | - | - |
| (Pillai et al., 2021) | Observational, cross-sectional | Out-patient | Descriptive | Activity, sleep apnea | 18 (44 %)  69.50 | - | - |
| (Qiao et al., 2020) | Observational, cross-sectional | Out-patient | Descriptive | Speech | 20 (65 %)  72.00 | - | - |
| (Qi et al., 2023) | Observational, cross-sectional | Out-patient | Descriptive and diagnostic | Graphomotor | - | 34 (56 %)  77.15 | - |
| (Richards et al., 2021) | Observational, cross-sectional | Care facilities and home | Descriptive | Sleep | - | 21 | - |
| (Robens et al., 2019) | Observational, cross-sectional | Out-patient | Descriptive | Graphomotor | 64 (55 %)  67.90 | - | - |
| (Robin et al., 2023) | Randomized controlled trial (longitudinal observations) | Out-patient | Descriptive | Speech | - | - | 130 (58 %)  69.19 |
| (Robin et al., 2021) | Observational, longitudinal study | Community and out-patient | Descriptive | Speech | - | - | 14 (50 %)  76.10 |
| (Schröter et al., 2003) | Observational, cross-sectional | Out-patient | Descriptive | Graphomotor | 39 (59 %)  60.60 | 13  - | - |
| (Serra-Ano et al., 2019) | Observational, cross-sectional | Out-patient | Descriptive | Gait | - | 18  76.78 | - |
| (Shim & Shin, 2022) | Observational, cross-sectional | Out-patient | Descriptive | Blood pressure | 59 (71 %)  73.95 | - | - |
| (Skirrow et al., 2024) | Observational, cross-sectional | Out-patient | Feasibility | Speech | 13 (38 %)  75.00 | - | 46 (48 %)  68.93 |
| (Sun et al., 2024)^a^ | Observational, longitudinal cohort study | Community | Incidence | Activity | - | - | - |
| (Svetnik et al., 2021) | Randomized controlled trial | Out-patient | Descriptive | Activity | - | 226  - | - |
| (Tian et al., 2021)^j^ | Observational, longitudinal cohort study | Community | Incidence | Activity | - | - | 64  - |
| (Toosizadeh et al., 2019)^g^ | Observational, cross-sectional | Out-patient | Descriptive | Kinematics | 34 (53 %)  83.88 | - | - |
| (Varma & Watts, 2016)^d^ | Observational, cross-sectional | Community | Descriptive | Activity | - | 39 (28 %)  73.50 | - |
| (Varma et al., 2021)^d^ | Observational, cross-sectional | Community | Descriptive | Gait | - | - | - |
| (Vidoni et al., 2016) | Randomized cross-over trial | Out-Patient | Descriptive | Steps | 21 (43 %)  72.30 | - | - |
| (H. Wang et al., 2021) | Observational, cross-sectional | Out-patient | Descriptive | Blood pressure | - | 27 (44 %)  75.70 | - |
| (Y. Wang et al., 2023) | Observational, cross-sectional | Out-patient | Descriptive | Fine motor skills | 46 (67 %)  70.00 | - | - |
| (Wanigatunga et al., 2022)^j^ | Observational, cross-sectional | Community | Descriptive | Activity | 31  - | - | - |
| (Watts et al., 2016) | Observational, cross-sectional | Community | Descriptive | Activity | - | 33  72.73 | - |
| (Westerberg et al., 2010) | Observational, cross-sectional | Out-patient | Descriptive | Sleep | 10 (80 %)  71.10 | - | - |
| (Wu et al., 2023)^a^ | Observational, longitudinal cohort study | Community | Descriptive | Activity | - | - | - |
| (Xiu et al., 2022) | Observational, cross-sectional | Out-patient | Descriptive | Speech | 11 (64 %)  73.36 | - | - |
| (Yamada, Kobayashi, et al., 2022)^h^ | Observational, cross-sectional | Out-patient | Diagnostic | Graphomotor | - | - | - |
| (Yamada, Shinkawa, Kobayashi, Caggiano, et al., 2021)^h^ | Observational, cross-sectional | Out-patient | Diagnostic | Gait, speech, graphomotor | - | - | - |
| (Yamada, Shinkawa, Kobayashi, Nishimura, et al., 2021)^h^ | Observational, cross-sectional | Out-patient | Diagnostic | Speech | - | - | - |
| (Yamada et al., 2023)^h^ | Observational, cross-sectional | Out-patient | Diagnostic | Speech | 46 (39 %)  73.80 | - | - |
| (Yamada, Shinkawa, et al., 2022)^h^ | Observational, cross-sectional | Out-patient | Diagnostic | Speech | - | - | - |
| (Yang et al., 2022) | Observational, cross-sectional | Out-patient | Descriptive | Kinematics | - | 106  - | - |
| (Yan & Dick, 2006) | Observational, cross-sectional | Out-patient | Descriptive | Graphomotor | 29 (45 %)  73.90 | - | - |
| (Yesavage et al., 1998) | Observational, longitudinal cohort study | Out-patient | Descriptive | Activity | - | 27  - | - |
| (Yesavage et al., 2002) | Observational, longitudinal cohort study | Out-patient | Descriptive | Rest/activity | - | 15  - | - |
| (N.-Y. Y. Yu & Chang, 2019) | Observational, cross-sectional | Out-patient | Descriptive | Graphomotor | 14 (29 %)  74.90 | - | - |
| (N. Y. Yu & Chang, 2016)^i^ | Observational, cross-sectional | Out-patient | Descriptive | Graphomotor | - | - | - |
| (Zhang et al., 2023) | Observational, cross-sectional | Out-patient | Descriptive | Graphomotor | 98 (57 %)  72.47 | - | - |

- Indicates that the demographic data variable is not applicable to the study

**Supplementary Table 2.** An overview of digital health technologies applied in the included studies, the software/algorithm used and details regarding the digital biomarkers obtained.

| **First author, year** | **Digital health technology** | **Digital health technology, specified** | **Analysis software/algorithm** | **Digital biomarker details** |
| --- | --- | --- | --- | --- |
| (André et al., 2019) | Actigraph | MotionWatch 8 wrist-worn triaxial actigraph (CamNTech Ltd, Cambridge, UK) | MotionWare software | Sleep fragmentation |
| (Balouch et al., 2022) | Actigraph | Model not stated (CamNtech Ltd) | Not stated | Sleep latency (aSleepLat), sleep period time (aSPT; interval between sleep start and wake time), total sleep time (aTST), sleep efficiency (aSE; actigraphy measured total sleep time/actigraphy measured time in bed), number of awakenings (aNAW), percentage of minutes immobile (aImmo; number of minutes immobile/aSPT), and mean length of immobility periods |
| (Basta et al., 2019) | Actigraph | Actilife v6.9.5, GT3XP model (Pensacola, FL, USA) | ActiLife 6 software | night sleep efficiency (SE), night sleep onset latency (SOL), night total sleep time (TST), night time in bed (TiB), night wake time after sleep onset (WASO), number of awakenings during the night, average duration of awakenings in the night, 24-hr (night-time and daytime) sleep time (24-hr TST) and 24-hr (night-time and daytime) time in bed (24-hr TiB) |
| (Beltrami et al., 2018) | Audio recorder | Olympus Linear PCM Recorder LS-5 | Transcriber software package | Linguistic and acoustic features |
| (Buchman et al., 2012) | Actigraph | Actical; Mini Mitter, Bend, OR | Not described | Total physical activity (activity counts) |
| (Buchman et al., 2020) | Accelerometer | Dynaport MT; McRoberts B.V., the Netherlands | Proprietary (MATLAB) analyses | Timed Up and Go (TUG), Standing Posture |
| (Buegler et al., 2020) | Tablet (Mobile app) | Altoida iADL test | Altoida application | Virtual placement of hidden objects that are found again by the user using a tablet or mobile phone |
| (Callow et al., 2024) | Actigraph | Actiwatch-2, Philips Respironics, Bend, OR | Actiware Software (v. 6.0.9) | total sleep time (TST; the number of minutes slept while in bed), and sleep efficiency (SE; the proportion of time in bed asleep, %) |
| (Callow et al., 2025) | Actigraph | Actiwatch-2, Philips Respironics, Bend, OR | Actiware Software (v. 6.0.9) | total sleep time (TST; the number of minutes slept while in bed), and sleep efficiency (SE; the proportion of time in bed asleep, %) |
| (Camargos et al., 2014) | Actigraph | Actiwatch Respironics, Inc. (Mini-Mitter, Bend, OR) | Actiware, version 5.59.0015, 2010 | Nighttime waking after sleep onset (WASO) (in minutes) during the nocturnal and prior to final waking. Number of nighttime awakenings (Awakenings) during the nocturnal period, after sleep onset, and prior to the awakening. Daytime total sleep time (DTST), in minutes, during the daytime period. Number of daytime naps (NAPS) during the daytime period. Nighttime percent sleep (%Sleep) during the nocturnal period, after sleep onset until the final awakening. |
| (Cavuoto et al., 2019) | Actigraph | Actiwatch 2 Mini-Mitter (Phillips-Respironics, OR, USA) | Actiware 6 | Wake after sleep onset (WASO, total minutes scored as wake following initial onset of sleep), sleep onset latency (minutes taken to fall asleep), total sleep time (total duration of the time in bed period spent asleep), sleep efficiency (SE; the proportion of time spent asleep compared to time spent in bed), sleep fragmentation index (a measure of mobility and short sleep bouts, it is the sum of percent mobile and percent of one minute immobile bouts divided by the number of immobile bouts for the sleep interval), and time in bed (TIB; the time in between Bed-time and Rise-time) |
| (Chen et al., 2020) | Inertial sensor unit | G-WALK (BTS Bioengineering Corp MA, United States) | BTS G-Studio (Copyright BTS Bioengineering S. p. A.) | Total timed-up and go (TUG) duration and each time of the TUG component were recorded: time to stand, time to turn around midway, time to turn around to reach the chair, and time to sit down in the chair. |
| (Corbi & Burgos, 2022) | Actigraph | AX3 device from Axivity, Ltd. | Estimation of Stationary Sleep Segments (ESS) algorithm | Wake After Sleep on Set (WASoS) and Total Sleep Time (TST) |
| (Costa et al., 2016) | Inertial sensor unit | Five kinetic sensing modules harboring 8051 microprocessor embedded in CC2530 Texas Instrument SoC (System on Chip) [20] and an inertial measurement unit MPU6000 (triaxial accelerometer and gyroscope) | Proprietary | Kinematic parameters are as follows: total displacement on the transverse plane (cm); maximal displacement (cm) with respect to the origin; mean distance (cm) with respect to the origin on the transverse plane, dispersion radius (average distances relative to average point), maximal and mean linear velocity (cm/s); positioning (cm) on X-axis (maximal, mean, and range) and Y-axis (maximal, mean, and range); roll angle (degrees) (maximal, minimum, and mean); and pitch angle (degrees) (maximal, minimum and mean). |
| (Egas-López et al., 2022) | Audio recorder | Olympus Digital Voice Recorder (WS-311M) | Proprietary |  |
| (El Haj et al., 2022) | Eye-tracker (pupillometer) | Pupil Lab | Pupil Capture software |  |
| (García-Gutiérrez et al., 2023) | Audio recorder (tablet application) | acceXible platform app | OpenSmile (v2.4.2) | Acoustic speech parameters (88 in total) |
| (García-Gutiérrez et al., 2024) | Audio recorder (tablet application) | acceXible platform app | OpenSMILE | Relevant acoustic parameters for detecting physiological changes in voice production |
| (Ghosal et al., 2022) | Actigraph | GT3x+ (Pensacola FL; Actigraph, 2012; 30 Hz sampling rate) | Proprietary | Further elaboration of analytical aspects of time-distributed data |
| (Gillain et al., 2016) | Accelerometer | Locometrix | Proprietary (MATLAB) | Gait speed, measured using a timing line and expressed in meters/second. Stride frequency or number of cycles per second (Hertz), calculated from the cranio-caudal acceleration following application of a Fourier transform. Stride length, deduced from the equation speed frequency 9 stride length and expressed in meters. Regularity, measured by the similarity (in terms of duration and amplitude) of the shape of cranio-caudal acceleration curves from steps and strides. This parameter is expressed in absolute value. Symmetry, defined as the similarity (in terms of duration and amplitude) of the shape of cranio-caudal acceleration curves when focusing on the right and left steps. This parameter is expressed in absolute value. |
| (Gillain et al., 2009) | Accelerometer | Locometrix | Proprietary | Gait speed measured using a timing line and expressed in metres per second the stride frequency or number of cycles per second (Hertz) is calculated from the craniocaudal acceleration following application of a Fourier transform stride length is deduced from the equation (speed = frequency stride length), stride regularity measures the similarity (in terms of duration and amplitude) of the shape of craniocaudal acceleration curves from one step to another; stride symmetry: the similarity (in terms of duration and amplitude) of the shape of craniocaudal acceleration curves when comparing right and left strides, specifically. |
| (Gosztolya et al., 2019) | Audio recorder | - | Proprietary | Acoustic parameters |
| (Gramkow et al., 2024) | Pupillometer | PLR-3000, NeurOptics | Not applicable | Baseline pre-stimulus pupil diameter, peak constriction pupil diameter, delta change between baseline and peak pupil diameter, latency, average constriction velocity, maximum constriction velocity, average dilation velocity, and time to reach 75% of baseline value after peak constriction. |
| (Granholm et al., 2017) | Pupillometer | NeurOptics PLR-200 (Irvine, CA; accuracy 0.1 mm, NeurOptics, 2010) | NA | Pupil dilation in response to cognitive load |
| (Guarnieri et al., 2020) | Actigraph | Fitbit Flex | Proprietary | Total sleep time, Wake after sleep onset, Sleep efficiency, acrophase, nadir, amplitude, mesor, period, circadian quotient |
| (Hajjar et al., 2023) | Audio recorder | iPod | Proprietary | Acoustic and lexical-semantic parameters |
| (Harms et al., 2022) | Accelerometer, tactile sensor | Tablet application | Altoida ML | Virtual placement of hidden objects that are found again by the user using a tablet or mobile phone |
| (Hatfield et al., 2004) | Actigraph | Actiwatch; Cambridge Neurotechnology, Cambridge, UK | Clocklab software (Actmetrix, Evanston, IL, USA) | Inter-daily stability (IS) is a measure of stability across days, while intra-daily variability (IV) reflects the relative consolidation/fractionation within days based on how many transitions occur between activity and rest. The third variable, rhythm amplitude (RA), reflects the difference in activity level between the 10 most active and least active hours in the day (L5) |
| (Hegedűs et al., 2024) | Audio recorder | Mobile phone (not stated) | Speech-Gap | Several acoustic speech parameters |
| (Huang et al., 2022) | Inertial measurement unit | JiBuEn gait analysis system | Proprietary | Stride speed, cadence, and heel strike angles came from the Free Walking test. TUG test measures in seconds, which is the time needed to rise from a chair, walk three meters, turn around, and return to a seated position at a faster speed. |
| (Hu et al., 2009) | Actigraph | Actiwatch (Mini Mitter/Respironics) | Proprietary | Scale-invariance of activity patterns |
| (Ishikawa et al., 2019) | Tablet | Digital drawing tablet (Wacom Cintiq Pro 16) | Proprietary | Numbers of segments were counted to characterize the handwriting behaviors. As kinematic parameters, velocity (m/s), acceleration (m/s2 ), and jerk (m/s3 ) of the pen-tip movements over the 2D coordinates (x, y) on the tablet surface were computed. The durations (s) of on-screen and in-air stylus pen movements were considered as timing parameters. |
| (Iwata et al., 2013) | Accelerometer | HJA-350IT; Omron, Kyoto, Japan | Omron (Bi-link version 1.0, Omron) | Amount of exercise performed carried out during specific time intervals. |
| (Jeon et al., 2023) | Accelerometer | SHIMMER | Proprietary | 35 features, including gait parameters and temporal features |
| (Kaneshwaran et al., 2019) | Actigraph | Actical, Phillips Respironics, Bend, OR | Proprietary | Sleep fragmentation |
| (Kawasaki et al., 2020) | Pupillometer | Handheld monocular pupillometer (Neurolight, IDMed, Marseilles) | Proprietary | Pupillary responses to blue and red light stimuli |
| (Khosroazad et al., 2023) | Sensor mattress and actigraph | piezo-resistive pressure sensors to record movement and respiratory signals using 54Hz average sampling rate. Actigraph Actiwatch 2 (Philips Respironics, Philips Actiware 6: v.6.0.9) | Proprietary | Sleep movement (SM) bouts (periodicity circa) and respiration |
| (Khou et al., 2018) | Actigraph | Actigraph GT3X+ | ActiLife V6.10.4 software and the Cole Kripke algorithm | Sleep onset latency, TST, TIB, SE ((TST/TIB) 100), WASO (amount of time spent awake after falling asleep), and average number of awakenings. |
| (Kim et al., 2021) | Actigraph | Actiwatch 2; Philips Respironics, Murrysville, PA, USA | Actiware-Sleep Software (version 6.0.2, Philips Respironics, Murrysville, PA, USA) | Sleep parameters from actigraphy |
| (Kimura et al., 2023) | Accelerometer | SilmeeTM W20, TDK Corporation, Tokyo, Japan | Not stated | Sleep parameters included the total sleep time (TST), sleep efficiency, waking frequency, and time awake after sleep onset (WASO). Furthermore, naptime was calculated from the time spent resting without moving during the daytime. In addition, physical activity was assessed and steps were defined as the frequency range of 23 Hertz of acceleration. |
| (Kobayashi et al., 2022) | Tablet | Wacom Cintiq Pro 16; sampling rate: 180 Hz; pen pressure levels: 8,192; pen inclination resolution: 1 degree; | Proprietary | The features consisted of 17 motion-related features (six related to speed and acceleration, five related to pen pressure, and six related to pen posture) and five pause-related features. |
| (König et al., 2017) | Accelerometer | CE-marked accelerometer research prototype (developed by Philips Research Laboratories Europe), wrist-worn | In-house developed algorithm | Walking speed, steps, and step variance |
| (Konig et al., 2017) | Audio recorder | SmartLav wearable microphone from the company Rode | Software tool PRAAT and self-developed tools | Vocal markers, mostly acoustic features, pauses |
| (Kuhlmei et al., 2013) | Actigraph | Actiwatch Mini, Cambridge Neurotechnology | Commercial software (not specified) | Activity profiles |
| (Lassila et al., 2018) | Blood pressure and heart rate monitor | Holter device (Cardioline walk200b, Cardioline S.p.A., Milan, Italy) | Self-developed algorithm | 24h estimation of heart rate and blood pressure |
| (Lazarou et al., 2023) | EEG headband, accelerometer | MUSE 2 device (InterAxon Inc., Toronto, ON, Canada) | MATLAB toolboxes | Power of the brain waves (alpha, beta, and theta) from four different electrodes (TP9, TP10, AF7 and AF8) |
| (Leger et al., 2016) | Actigraph | Motionwatch 8 (MW8, Camntech, Cambridge, UK) | Motion Ware software 2.5 (Camntech Ltd) | Sleep was scored when the total activity count (A) was equal to or less than the activity threshold setting according to the following formula: A = an2(1/25) + an1(1/5) + a + a1(1/5) + a2(1/25); where an2 and an1 were the activity counts from the prior 2 min, and a1 and a2 the subsequent 2 min |
| (K. Li et al., 2023) | Tablet | Not specified | Self-developed | Mean drawing speed, maximum drawing speed, minimum drawing speed, and drawing speed variability. total task time, start writing time, total drawing time, total pause time, mean pause time, maximum pause time, variability of pause time, drawing pause rate, and fingertip retention rate. |
| (Lim et al., 2013) | Actigraph | Actical (Phillips Respironics, Bend, OR) | MATLAB algorithm | Sleep fragmentation |
| (P. Li et al., 2020) | Actigraph | Actical; Philips Respironics, Bend, OR, USA | MATLAB algorithm | Parametric cosine curve fitting and non-parametric analyses |
| (P. Li et al., 2023) | Actigraph | Actical; Philips Respironics, Bend, OR, USA | MATLAB algorithm | Daytime napping |
| (P. Li et al., 2018) | Actigraph | Actical, Philips Respironics, Bend, OR | MATLAB algorithm | Fractal regulation of activity |
| (Louzada et al., 2022) | Actigraph | ActTrust AT0503 actigraphs (Condor Instruments) | ActStudio software (version 1.0.5.3) | Main nocturnal sleep duration. Night time waking after sleep onset, number of awakenings during nocturnal sleep, daytime total sleep time, number of daytime naps |
| (Luboshitzky et al., 2001) | Actigraph | Mini-ACT, AMA-32, AMI, Ardsley, NJ | Actigraphic Scoring Analysis (ASA) software program | Sleep onset time, sleep wake-up time, sleep period time, true sleep time, sleep efficiency (SE), longest episode of continuous sleep, minutes of awakening during sleep, minutes of no activity |
| (Lu et al., 2018) | Actigraph | Actigraph wGT3x-BT accelerometer (Pensacola, Florida, USA) | ActiGraph algorithm available in the ActiLife software | Daily average VM cpm, percentage of wear-time spent in sedentary behavior and average VM cpm during sleeping hours. Sedentary behavior. Average length of sedentary bouts, total numbers of sedentary bouts, number of long sedentary bouts, numbers of sedentary breaks (defined as a period of 1 3 consecutive min when the accelerometer registered, number of long non-sedentary bouts. |
| (Lu et al., 2024) | Actigraph | Actigraph wGT3x-BT accelerometer (Pensacola, Florida, USA) | cosinor and nparACT packages in R | Mesor, amplitude, acrophase, interdaily stability, intradaily variability, L5, M10, M10 onset time, relative amplitude |
| (Lysen et al., 2020) | Actigraph | ActiWatch model AW4, Cambridge Technology Ltd | Not stated | Intradaily variability, interdaily stability, L5 onset |
| (Maquet et al., 2010) | Accelerometer | Locometrix; Centaure-Metrix, Evry Cedex, France) includes an acceleration sensor, a recording device, and a computer program for processing the acceleration signals. | analyzed by software developed in the MATLAB 5 environment (Mathworks, France). | Comfortable walking speed, stride frequency, stride length, step symmetry, stride regularity, number of stops during walking in simple and dual tasks |
| (Mc Ardle et al., 2023) | Actigraph | Axivity AX3, York, UK | brms package for R statistical software | daily step count and pattern, mean bout length, alpha |
| (Mc Ardle, Del Din, Donaghy, et al., 2020) | Actigraph | Axivity AX3, York, UK | MATLAB | Total walk time, total steps and total bouts, mean length of walking bouts, and alpha, variability of bout length between walking bouts |
| (Mc Ardle, Del Din, Galna, et al., 2020) | Actigraph | AX3, Axivity, York, UK; dimensions 23.0 mm x 32.5 mm x 7.6 mm; 11gms; 512Mb memory; 100 Hz, 10-bit resolution, 8 g range | MATLAB | Pace (step velocity, step length, step time variability), variability (swing time variability, stance time variability, step velocity variability, step length variability), rhythm (step time, swing time, stance time), asymmetry (step time asymmetry, swing time asymmetry, stance time asymmetry) and postural control (step length asymmetry) |
| (Ardle et al., 2018) | Actigraph | Axivity AX3; Axivity, York, UK; Dimensions: 23.0mm x32.5mm x7.6mm, weight 9g) | Not stated | Variability was estimated from the standard deviation between all steps. Asymmetry was determined as the absolute difference between left and right steps |
| (Meier et al., 2021) | Tablet | Altoida DNS | Altoida DNS | Gait, touch pressure, walk path, tremor |
| (Meilan et al., 2017) | Audio recorder | Not stated | Praat 5.1.4231 voice-analysis program | Non-periodicity, interruption, amplitude disturbance, and resonance and noise disturbance in the voice signal |
| (Moghadami et al., 2021) | Eye-tracker | SMI eye-tracker (Senso Motoric Instruments) | Be Gaze (V. 3.7) software | Areas with many fixations |
| (Müller et al., 2017) | Digitizing tablet | Windows Surface Pro 4 digitizer with a handheld stylus pen | Not stated | Time-in-air, time-on-surface, total-time |
| (Muurling et al., 2020) | Actigraph | ActiGraph wGT3X-BT accelerometers (ActiGraph LCC, Pensacola, FL) | ActiLife 6 application (version v6.11.4) and custom MATLAB script. | Mean stance time, stride time, swing time, step length and velocity, step frequency, and stance time, stride time, swing time, and step length variability |
| (Muurling et al., 2024) | Several (tablet, actigraph, accelerometer, EEG headband) | Altoida app, Axivity AX3 (actigraph), Fitbit Charge 3 (accelerometer), Dreem (EEG headband) | Not stated | Activity, sleep, heart rate |
| (Nasrolahzadeh et al., 2018) | Audio recorder | Not stated | Not stated | Different acoustic parameters |
| (Park et al., 2021) | Actigraph | Actiwatch 2 (Philips Respironics, Murrysville, PA, USA) | Actiware, version 6.0.9 | Bedtime, wake-up time, total sleep time, sleep onset latency, sleep efficiency, and wake time after sleep onset. Cosinor and nonparametric analyses |
| (Perez-Valero et al., 2022) | Wearable EEG | Versatile wireless wearable system by Bitbrain | Not stated | Various EEG features |
| (Petrillo et al., 2023) | Gyroscope | Tri-axial wearable gyroscopes (sample frequency = 100 Hz, BioSensics LLC, Boston, MA, USA) | Not stated | Angular velocity |
| (Pillai et al., 2021) | Actigraph | Motionlogger Micro Watch by Ambulatory Monitoring, Inc | Cole-Kripke algorithm | F-ratio, amplitude, mesor and acrophase. Sleep regularity index interdaily stability, total sleep time (TST), nocturnal awakenings, sleep efficiency, wakefulness after sleep onset, sleep fragmentation index |
| (Qiao et al., 2020) | Audio recorder | Not stated | ASR software V1.3 (China Software Copyright | Several speech parameters |
| (Qi et al., 2023) | Digital pen | dot-matrix pen (TSTUDY, China) | Not stated | 35 handwriting characteristics |
| (Richards et al., 2021) | Actigraph | Micro-Mini Motionlogger Actigraph (Ambulatory Monitoring Inc., Ardsley, NY) | Ambulatory Monitoring software | Estimate of the true sleep period, sleep minutes, wake minutes, sleep fragmentation index |
| (Robens et al., 2019) | Digitizing tablet | Microsoft Surface Pro 3 digitizer. | Not stated | Several graphomotor parameters |
| (Robin et al., 2023) | Audio recorder | tablet computers (Virgil platform, WCG MedAvante-ProPhase, Hamilton NJ) | Python-based custom algorithm | Acoustic parameters |
| (Robin et al., 2021) | Audio recorder | iPad, WinterLight assessment | WinterLight labs pipeline (Python) | Acoustic (e.g., properties of the sound wave, speech rate, number of pauses) |
| (Schröter et al., 2003) | Digitizing tablet | WACOM-IV digitizing tablet and a pressure-sensitive stylus. | CS 4.3 software | Mean peak velocity, the standard deviation of the intraindividual velocity profile, writing frequency |
| (Serra-Ano et al., 2019) | Gyroscope, accelerometer | HighPerformance 6-Axis MEMS MotionTracking composed of 3-axis gyroscope (gyro), 3-axis accelerometer (acc), and a Digital Motion Processor (TDK- ICM-20689) at 100 Hz. | Custom software in Python | Postural control, gait kinematics |
| (Shim & Shin, 2022) | Blood pressure monitor | TM-2430; A&D, Tokyo, Japan | Not stated | Average systolic blood pressure (sBP) and diastolic BP for the daytime, nighttime, and 24-hour periods. Mean sBP, mean dBP, and standard deviations of both were collected over 24 hours. |
| (Skirrow et al., 2024) | Smartphone | Novoic app | Novoic app, proprietary | Recall of automated story recall task which extracts features from speech |
| (Sun et al., 2024) | Actigraph | Actical, Philips Respironics, Bend, OR, US | Not stated | Physical activity and fragmentation of activity, inter-daily stability, intra-daily variability |
| (Svetnik et al., 2021) | Actigraph | Garmin vivosmart HR device that measures movement using a tri-axial accelerometer | Proprietary | 24/7 timing information (onset, end, duration) regarding periods of putative light sleep, deep sleep and wake, as well as other daytime activity (steps, naps) and heart rate |
| (Tian et al., 2021) | Actigraph | ActiHeart; CamNtech | ActiHeart, version 4.0.32 | Active-to sedentary transition probability |
| (Toosizadeh et al., 2019) | Gyroscope, accelerometer | BioSensics LLC, Boston, MA, USA | In-home developed MATLAB script | Elbow flexion: agility (speed, rise time, and flexion number) |
| (Varma & Watts, 2016) | Actigraph | GT3x+ (Pensacola FL; Actigraph, 2012; 30 Hz sampling rate) | Custom developed algorithm | Activity phase of the daily diurnal cycle, accelerometry-derived measures of physical activity |
| (Varma et al., 2021) | Actigraph | Actigraph GT3X+ (Pensacola FL; Actigraph, 2012) | Custom developed algorithm | Walking, lying, standing, and sitting activity. Walking bouts of at least 60 seconds were evaluated to generate 55 gait variables within five domains: amplitude, pace, rhythm, symmetry, and variability. |
| (Vidoni et al., 2016) | Actigraph | Zip (FitBit Inc., San Francisco, CA) | Proprietary, open-source software (Cran-R v3.1.1, | Step counts |
| (H. Wang et al., 2021) | Blood pressure monitor | 24 h ambulatory BP monitors (MobilO-Graph, I.E.M., Stolberg, Germany) | Not stated | Means of the 24 h, daytime, and night-time ambulatory systolic and diastolic blood pressure were calculated. Standard deviation and coefficient variation were used as the measures of variability. Circadian blood presusure rhythm was assessed by dipping pattern of systolic and diastolic BP, reverse dipping, reduced dipping, normal dipping, and extreme dipping |
| (Y. Wang et al., 2023) | Tablet | human-computer interaction | Not stated | Execution time for motor skill |
| (Wanigatunga et al., 2022) | Actigraph | GT9X Actigraph (Pensacola, FL) | ActiLife software and (ver. 6.13.4) and ARCTOOLS package in R | Activity counts per day, active minutes per day, activity fragmentation |
| (Watts et al., 2016) | Actigraph | Actigraph GT3X+ (Pensacola, FL) | Not stated | Vector magnitude, mean and variability of activity |
| (Westerberg et al., 2010) | Actigraph | Wrist-worn, model not stated | Not stated | Not stated |
| (Wu et al., 2023) | Actigraph | Not stated | Not stated | Rest fragmentation |
| (Xiu et al., 2022) | Audio recorder | Microphone (Sennheiser e 835) with digital recorder (TASCAM DR40X) | Praat | F0 values, formant frequencies (F1, F2 and F3) |
| (Yamada, Kobayashi, et al., 2022) | Digitizing tablet | Wacom Cintiq Pro 16; sampling rate: 180 Hz; pen pressure levels: 8,192; pen inclination resolution: 1 degree; screen size: 345 194 mm (2560 1440 pixels | Not stated | Drawing speed, variability and non-smoothness, pressure variability and non-smoothness, mean pause. duration between drawing, the pause drawing duration ratio, and adjusted total duration |
| (Yamada, Shinkawa, Kobayashi, Caggiano, et al., 2021) | Motion sensor, audio recorder and digitizing tablet | OptiTrack Flex13, sampled at 120 Hz using OptiTrack Motive software 2.1.0 Beta 1 (NaturalPoint, Inc, Corvallis, OR, USA). tablet device (iPad Air 2), iPads internal microphone (core audio format, 44,100 Hz, 16-bit). | Not stated | Gait speed and step stride length, rhythm, variability, left-right asymmetry, and postural control. Mel-frequency cepstral coefficients, pitch variability, proportion of pause duration, proportion of mistakes in both calculation tasks, Honoré's statistic. Drawing speed, pressure-related, time-related, time duration between nodes |
| (Yamada, Shinkawa, Kobayashi, Nishimura, et al., 2021) | Audio recorder | internal microphone of the iPad (core audio format, 44,100 Hz, stereo, 16-bit) | Not stated | Mel frequency cepstral coefficients, the first three formant frequencies (F1 F3), jitter (local, RAP, PPQ5, DDP), and shimmer (local, APQ3, APQ5, APQ11, DDA) |
| (Yamada et al., 2023) | Audio recorder | iPad Air tablet | we used the Python (version 3.8) audio processing libraries librosa (version 0.8.0) (McFee et al., 2015) for MFCCs, and Signal_Analysis (version 0.1.26) | Jitter, shimmer, and mel-frequency cepstral coefficients |
| (Yamada, Shinkawa, et al., 2022) | Audio recorder | iPad Air 2 Tablet | Not stated | Jitter and shimmer, variances of first-order derivatives of the first 12 Mel frequency cepstral coefficients |
| (Yang et al., 2022) | Accelerometer | Microsoft Kinect sensor (Microsoft Corp., Redmond, WA, USA) | Microsoft Kinect Software Development Kit (SDK) 2.0 (Microsoft Corp., Redmond, WA, USA) | Spatial location of 15 human joints in three dimensions |
| (Yan & Dick, 2006) | Digitizing tablet | WACOM digitizer-tablet (UD-1218-R) and a hand-held stylus pen | Custom developed algorithm | Various graphomotor parameters |
| (Yesavage et al., 1998) | Actigraph | Ambulatory Monitoring Systems, Inc., Ardsley, NY 10502 | ACTION 1.3 (Ambulatory Monitoring Systems). | Amount of time spent in bed during the night sleep period, time to sleep onset at the beginning of the sleep period, total time spent in sleep during the sleep period, the amount of time awake after sleep onset, and sleep efficiency |
| (Yesavage et al., 2002) | Actigraph | Ambulatory Monitoring Systems, Inc., Ardsley, NY | ACTION software version 1.32 | Amplitude , sleep efficiency |
| (N.-Y. Y. Yu & Chang, 2019) | Digitizing tablet | Wacom Intuos 5, Japan | Not stated | Movement fluency, handwriting accuracy |
| (N. Y. Yu & Chang, 2016) | Digitizing tablet | Wacom Intuos 5, Japan | Not stated | Movement speed, various graphomotor parameters for drawing task. |
| (Zhang et al., 2023) | Digitizing tablet | PH-1820-A; PendoTech | Custom phyton script | Time, writing pressure, jerk |

**Search strings**

**MEDLINE**

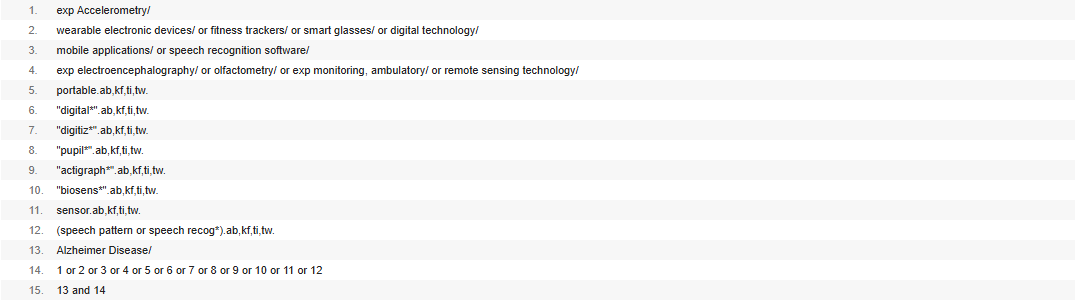

**Web of Science**

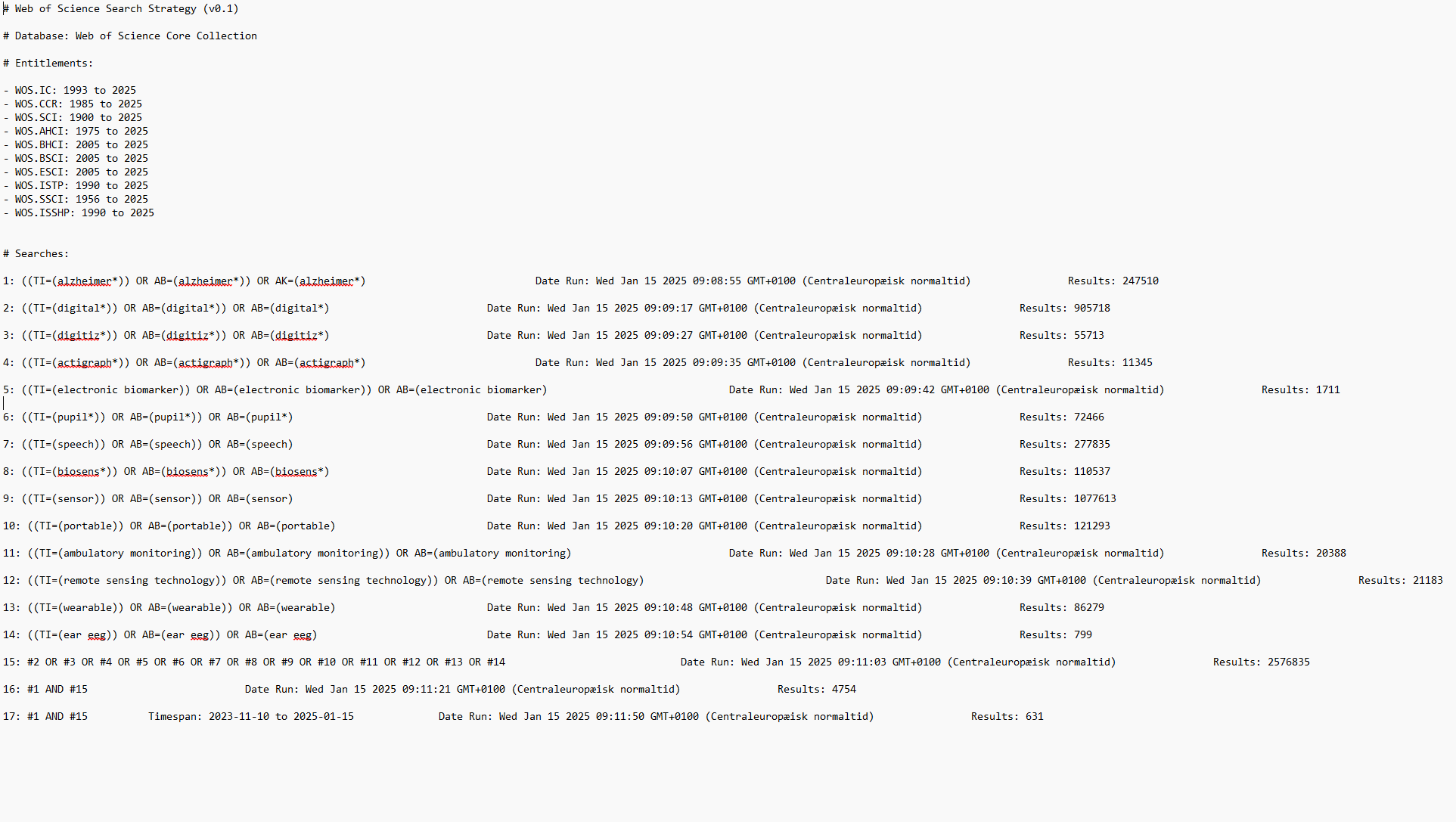

**EMBASE**

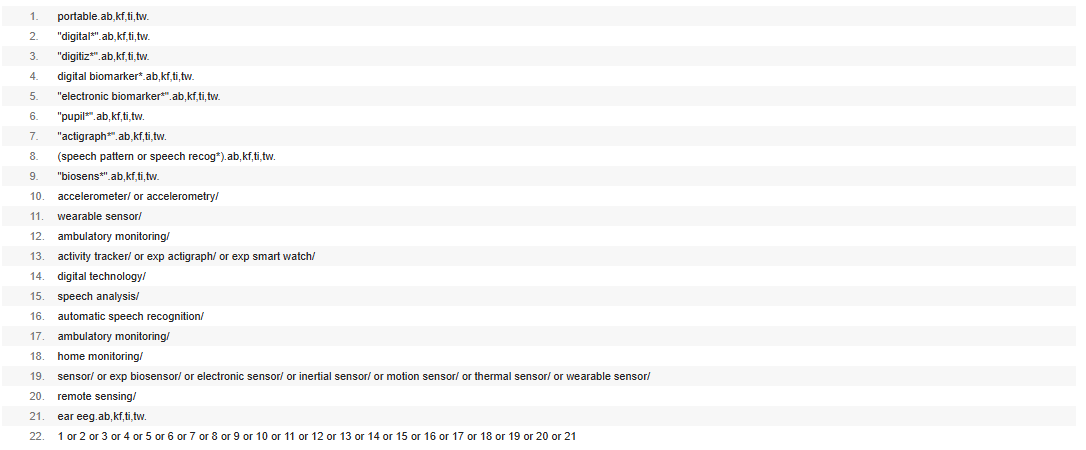
